# Analysis of immunization time, amplitude, and adverse events of seven different vaccines against SARS-CoV-2 across four different countries

**DOI:** 10.1101/2022.03.11.22272153

**Authors:** Maria Elena Romero-Ibarguengoitia, Arnulfo González-Cantú, Chiara Pozzi, Riccardo Levi, Maximiliano Mollura, Riccardo Sarti, Miguel Angel Sanz-Sánchez, Diego Rivera-Salinas, Yodira Guadalupe Hernández-Ruíz, Ana Gabriela Armendariz-Vázquez, Gerardo Francisco Del Rio-Parra, Irene Antonieta Barco-Flores, Rosalinda González-Facio, Elena Azzolini, Riccardo Barbieri, Alessandro Rodrigo de Azevedo Dias, Milton Henriques-Guimaraes, Alessandra Bastos-Borges, Cecilia Acciardi, Graciela Paez-Bo, Mauro Martins Teixeira, Maria Rescigno

## Abstract

**Background:** Scarce information exists in relation to the comparison of seroconversion and adverse events following immunization (AEFI) with different SARS-CoV-2 vaccines. Our aim was to correlate the magnitude of the antibody response to vaccination with previous clinical conditions and AEFI.

**Methods:** A multicentric comparative study where SARS-CoV-2 spike 1-2 IgG antibodies IgG titers were measured at baseline, 21-28 days after the first and second dose (when applicable) of the following vaccines: BNT162b2 mRNA, mRNA-1273, Gam-COVID-Vac, Coronavac, ChAdOx1-S, Ad5-nCoV and Ad26.COV2. Mixed model and Poisson generalized linear models were performed.

**Results:** We recruited 1867 subjects [52 (SD 16.8) years old, 52% men]. All vaccines enhanced anti-S1 and anti-S2 IgG antibodies over time (p<0.01). The highest increase after the first and second dose was observed in mRNA-1273 (p<0.001). There was an effect of previous SARS-CoV-2 infection; and an interaction of age with SARS-CoV-2, Gam-COVID-Vac and ChAdOx1-S (p<0.01). There was a negative correlation of Severe or Systemic AEFI (AEs) of naïve SARS-CoV-2 subjects with age and sex (p<0.001); a positive interaction between the delta of antibodies with Gam-COVID-Vac (p=0.002). Coronavac, Gam-COVID-Vac and ChAdOx1-S had less AEs compared to BNT162b (p<0.01). mRNA-1273 had a higher number of AEFIs. The delta of the antibodies showed an association with AEFIs in previously infected individuals (p<0.001).

**Conclusions:** The magnitude of seroconversion is predicted by age, vaccine type and SARS-CoV-2 exposure. AEs are correlated with age, sex, and vaccine type. The delta of the antibody response is positively correlated with AEs in patients previously exposed to SARS-CoV-2.

**Registration number:** NCT05228912

## Introduction

Covid-19 is a pneumonia-like disease caused by a coronavirus, named SARS-CoV-2. It is highly contagious, and the World Health Organization declared it in 2020 as a pandemic (1). SARS-CoV-2 causes COVID-19 that has a wide range of clinical presentations, from asymptomatic disease to severe acute respiratory distress syndrome and death(2). SARS-CoV-2 utilizes its surface spike glycoprotein to enter host cells. Each unit of the spike trimer contains an S1 and S2 subunit, with the N-terminal S1 subunit binding to the cellular angiotensin-converting enzyme 2 (ACE2) receptor through an internal receptor-binding domain (RBD)(3).

To prevent SARS-CoV-2 infection, 113 vaccines are being tested in human clinical trials, and 44 have reached the final step of testing(4). Based on the mechanism of action, vaccines can be clustered in four groups: 1) mRNA vaccines, for example, BNT162b2 mRNA and mRNA-1273. They use genetically engineered modified RNA to produce the spike protein that prompts an immune response safely. 2) Viral vector (adenovirus) vaccines, for example, ChAdOx1-S, Ad26.COV2, Ad5-nCoV, and Gam-COVID-Vac. They use a virus that has been genetically engineered so that it cannot cause disease but produces coronavirus proteins to safely generate an immune response. 3) Inactivated virus vaccines, for example, Coronavac, use a form of the virus that has been inactivated or weakened, so it does not cause disease but still generates an immune response. 4) Protein subunits vaccines, for example, NVX-CoV2373. It is a protein-based vaccine that uses harmless fragments of proteins or protein shells that mimic the SARS-CoV-2 to safely generate an immune response(5).

Currently BNT162b2 mRNA, mRNA-1273, ChAdOx1-S, Ad26.COV2, Ad5-nCoV, Gam-COVID-Vac, Coronavac, have been administered to people across 587 countries(4). However, no studies have compared using the same assay and time frame the effectiveness of seroconversion and incidence of adverse event events in response to vaccines in different countries in the same study.

This study aimed to correlate the magnitude of the antibody response after the first and second dose (if applicable) between BNT162b2 mRNA, mRNA-1273, ChAdOx1-S, Ad26.COV2, Ad5-nCoV, Gam-COVID-Vac, and Coronavac by measuring SARS-CoV-2 spike 1-2 IgG antibodies, using the same standardized assay, with previous clinical conditions and systemic and severe adverse events (AEs).

## Materials and Methods

This was a multicentric observational study of volunteers who received an approved complete scheme of BNT162b2 mRNA, mRNA-1273, ChAdOx1-S, Ad26.COV2, Ad5-nCoV, Gam-COVID-Vac, or Coronavac COVID-19 vaccine during 2021 in five hospital centers (Hospital Clinica Nova, Humanitas Clinical and Research Center, Fundacion San Francisco Xavier, Ternium Health Center in Rio, Hospital Municipal San Jose, Hospital Interzonal de Agudos San Felipe) from four different countries: Mexico, Italy, Brazil and Argentina. This study followed STROBE guidelines (6). The study was approved by each local Institutional Review Board and conducted per the Code of Ethics of the World Medical Association (Declaration of Helsinki) for experiments that involve humans.

The inclusion criteria were volunteers of both genders, any age, who consented to participate, planned to conclude the immunization regimen of any vaccine, and agreed to be followed up for the duration of the study. The following vaccines were available: BNT162b2 mRNA, mRNA-1273, ChAdOx1-S, Ad26.COV2, Ad5-nCoV, Gam-COVID-Vac, or Coronavac. The exclusion criteria were to have received any SARS-CoV-2 vaccine prior to study entry.

Each country’s Health System defined the vaccines available, the schedule, and dose assignation. On the vaccination day, the research team invited any subject who planned to receive any vaccine scheme, explained the project and asked to sign the informed consent. Inclusion-exclusion criteria were applied, and a plasma sample was collected. The baseline sample was taken before receiving the first dose of any SARS-CoV-2 vaccine (T0). The second (T1) and third samples (T2) were taken after 21 days (+/- 7 days) of the first and second dose, respectively.

At each visit, participants had to answer a questionnaire. The basal questionnaire aimed at obtaining patients’ medical history and previous SARS-CoV-2 infections. The questionnaires applied after the first and second dose of vaccines aimed at recognizing adverse events following immunization (AEFI)(7) and identifying a potential SARS-CoV-2 infection after receiving any vaccine dose. SARS-CoV-2 infection was also monitored by the epidemiology team through PCR testing, which informed the research team of any new infection.

### Primary Outcome, IgG determination

Our primary outcome was to correlate the magnitude of the antibody response to vaccination with previous clinical conditions and AEs. To determine the amount of specific anti-S1 and anti-S2 IgG antibodies against SARS-CoV-2 in plasma samples, the laboratory personnel used a chemiluminescence immunoassay (CLIA) developed by DiaSorin, which had a sensitivity of 97.4% (95% CI, 86.8-99.5) and a specificity of 98.5% (95% CI, 97.5-99.2). The results were reported as follows: <12.0 AU/ml was considered negative, 12.0 to 15.0 AU/ml was indeterminate, and > 15 AU/ml was positive. This kit is comparable with other commercial kits and has been used elsewhere (8–11).

The variables we analyzed were age, sex, personal medical history (for example, the presence of type 2 diabetes, hypertension, obstructive pulmonary disease, any heart condition, obesity, cancer, liver steatosis, any autoimmune disease), and confirmed SARS-CoV-2 (through nasal swab or serologic tests). The following AEs were considered of particular interest: fever (>37.5°C), adenopathy, diffuse rash, edema, facial paralysis, orthostatic hypotension, headache, arthralgia, myalgia, nausea, vomit, and diarrhea. Anti-S1 and anti-S2 IgG antibodies against SARS-CoV-2 were measured at baseline, 21-28 days post-first dose (S1), and 21-28 days post-second dose if applicable.

### Statistical Methods

The researchers reviewed the quality control and the anonymization of the database. Normality assumption was evaluated with the Shapiro-Wilk test or Kolmogorov. Descriptive statistics such as mean, standard deviation, median, interquartile range, frequencies, and percentages were computed. Kruskal-Wallis test with Dunn’s multiple comparisons test was performed for IgG comparisons. We performed a mixed model where the dependent variable was the delta of the antibodies. The personal variability and time were constructed as a random effect, whereas each type of vaccine, age, history of SARS-CoV-2, and the interaction of each vaccine with age were fixed effects. BNT162b mRNA was the reference vaccine group. For the analysis of AEs, the study population was divided into two separate cohorts by stratifying according to the history of SARS-CoV-2 infection (1050 Naïve SARS-CoV-2 subjects and 471 SARS-CoV-2 history subjects). Poisson generalized linear models (PGLM) were performed on both cohorts with the counts of AEs events after the second dose as the outcome variables. Included regressors were age, sex, body mass index, 1,000 AU/mL variation of IgG levels after the second dose compared to the baseline IgG levels, type of vaccine and the interactions between vaccines and IgG level variation. BNT162b mRNA was the reference vaccine group Missing completely at random values were analyzed through complete case analysis since missing antibody levels were less than 5% and we considered it incorrect to impute any AEFI. A sample size of 1870 patients was calculated, according to the primary aim, by using a mixed model formula with an alpha of 0.05, power of 90%, the effect size of 0.15, and k=7. The statistical programs used were R v.4.0.3 and Python v. 3.8.3. The analyses were two-tailed. A p-value less than 0.05 was statistically significant.

## Results

A total of 1867 patients were recruited from all countries: 1352 from Mexico, 42 from Italy, 260 from Brazil, and 213 from Argentina. The most frequent vaccine used was ChAdOx1-S in 666 subjects, Coronavac in 582, BNT162b2 mRNA in 289, Gam-COVID-Vac in 213, mRNA-1273 in 65, Ad26.COV2 in 31, and Ad5-nCoV in 19. The mean (SD) age was 52 (16.8) years, being statistically different across vaccine groups (p<0.05) as some vaccines were proposed to a particular age group. Fifty-two % of subjects were men, 559 (30%) had obesity, and 501 (26.8%) had hypertension. Table 1 shows the medical history of patients divided by vaccine type.

**Table 1:** General characteristics and medical history Table 1 shows the medical history and comparison between vaccine groups, p<0.05 was considered statistically significant.

|  | BNT162b2 mRNA (%) | mRNA-1273(%) | Gam-COVID-Vac (%) | Coronavac (%) | ChAdOx1-S (%) | Ad5-nCoV (%) | Ad26.COV2(%) | p-value |
| --- | --- | --- | --- | --- | --- | --- | --- | --- |
| Total | 289 | 65 | 200 | 582 | 666 | 19 | 31 |  |
| Obesity | 54(18.6) | 11(16.9) | 44(22) | 188(32.3) | 248(37.2) | 8(42.1) | 6(19.3) | <0.001 |
| Smoker | 37(12.8) | 10(15.3) | 27(13.5) | 60(10.3) | 38(5.7) | 2(10.5) | 2(6.4) | 0.001 |
| Hypertension | 36(12.4) | 2(3) | 77(38.5) | 116 (19.9) | 265(39.7) | 3(15.7) | 2(6.4) | <0.001 |
| Dyslipidemia | 27(9.3) | 3(4.6) | 55(27.5) | 76 (13.5) | 150 (22.5) | 1(5.2) | 1(3.2) | <0.001 |
| Type2 Diabetes | 13(4.4) | 1(1.5) | 32(16) | 78(13.4) | 164(24.6) | 0 | 0 | <0.001 |
| Asthma | 9 (3.1) | 3(4.6) | 6(3) | 13 (2.3) | 17 (2.5) | 0 | 0 | 0.79 |
| Other autoimmune disease such as thyroiditis or psoriasis | 9(3.1) | 0 | 4(2) | 29 (4.9) | 60 (9) | 1(5.2) | 1(3.2) | <0.001 |
| Rheumatoid arthritis | 7(2.4) | 1(1.5) | 16(8) | 11(1.8) | 56 (8.4) | 0 | 1(3.2) | <0.001 |
| Surgery in the last year | 6(2) | 0 | 6(3) | 26(4.4) | 45(6.7) | 3 (15) | 1(3.2) | 0.002 |
| Previous Cancer | 5(1.7) | 1(1.5) | 1(0.5) | 5(0.8) | 29(4.3) | 0 | 0 | 0.001 |
| NAFLD | 4 (1.3) | 0 | 7(3.5) | 27(4.6) | 21(3.1) | 2(10.5) | 0 | 0.042 |
| Immunosuppressive treatment | 4(1.3) | 0 | 3(1.5) | 6(1) | 11(1.6) | 0 | 0 | 0.86 |
| Other liver disease | 3 (1) | 0 | 10(5) | 10(1.7) | 10(1.5) | 0 | 0 | 0.02 |
| End stage renal disease | 2 (0.7) | 0 | 2(1) | 3(0.5) | 11(1.6) | 0 | 0 | 0.44 |
| Pregnancy | 2(0.7) | 1(1.5) | 0 | 3(0.5) | 0 | 1(5.2) | 0 | 0.005 |
| Cirrhosis | 2 (0.7) | 0 | 0 | 2(0.3) | 4 (0.6) | 0 | 0 | 0.88 |
| Pulmonar Obstructive Chronic Disease | 1(0.3) | 0 | 3(1.5) | 4(0.6) | 8 (1.2) | 0 | 0 | 0.68 |
| Coronary heart disease | 1(0.3) | 0 | 15(7.5) | 3(0.5) | 23(3.4) | 0 | 0 | <0.001 |
| Gout | 1(0.3) | 0 | 10(5) | 19(3.2) | 27 (4.05) | 0 | 0 | 0.02 |
| Active Cancer | 0 | 0 | 1(0.5) | 1(0.17) | 10(1.5) | 0 | 0 | 0.054 |
| Atrial Fibrillation | 0 | 0 | 13(6.5) | 6(1) | 36 ( 5.4) | 0 | 0 | 0.001 |
| Congestive heart failure | 0 | 0 | 18(9) | 1(0.2) | 12( 1.8) | 0 | 0 | 0.001 |
| Stroke | 0 | 0 | 2(1) | 1(0.2) | 13(1.9) | 0 | 0 | 0.01 |
| Trasplant | 0 | 0 | 0 | 3(0.5) | 5 (0.7) | 0 | 0 | 0.63 |
| Pregnancy | 2 | 1 | 0 | 3(0.5) | 0 | 1(5.2) | 0 | 0.05 |

### History of SARS-CoV-2

Positive history of SARS-CoV-2 infection was considered if the volunteer had a confirmatory swab or serological test. Additionally, it was considered if specific anti-S1 and anti-S2 IgG antibodies against SARS-CoV-2 in plasma samples were > 15 AU/ml at baseline. We had a total of 627 positive cases before vaccination, of which 165 (24.8%) received ChAdOx1-S, 246 (42.2%) Coronavac, 107 (37.1%) BNT162b2 mRNA, 55 (25.8%) Gam-COVID-Vac, 31 (47.6%) mRNA-1273, 18 (58%) Ad26.COV2, and 5 (26%) Ad5-nCoV.

### Antibody titers

All vaccines showed significant anti-S1 and anti-S2 IgG antibody changes with significant differences across vaccines and depending on SARS-CoV-2 history. In naïve patients, the highest increase [Median (IQR) AU/ml] after the first dose was observed in mRNA-1273 [175.5 (76.7)], then BNT162b mRNA [78.1(55)], and Ad5-nCoV [43.4(55.9)]. In subjects previously exposed to SARS-CoV-2, the highest increase after the first dose was observed in mRNA-1273 [5920(4705)], then BNT162b mRNA [2500(3080)], and ChAdOx1-S [1025(2489)].

In naïve patients, the highest increase [Median (IQR) AU/ml] after the second dose (T2) was observed in mRNA-1273 [1875 (1190)], then BNT162b mRNA [998 (1241)], and Gam-COVID-Vac [501 (55.9)], whereas in SARS-CoV-2 previously exposed subjects the highest increase after the second dose was observed in mRNA-1273 [4950(3060)], then Gam-COVID-Vac [3620 (5356)], and BNT162b mRNA [2630 (2672.5)]. Figure 1 and Figure S1 show the change in antibody levels through time, classified by vaccine type. Table 2 shows the antibody levels classified by vaccine type. The mixed model that showed a positive effect of SARS-CoV-2 previous infection (B=767.468, p<0.001), a positive interaction of age with previous SARS-CoV-2 infection (B=13.03, p=0.003), and an interaction of Gam-COVID-Vac and ChAdOx1-S with age (B=46.5, p<0.001; B=20.5; p=0.006). With respect to BNT162b mRNA, mRNA-1273 showed a higher change in antibody titers, while all the other vaccines had less change (p<0.01). We computed sex in the model, but there was no significant correlation, so we eliminated it from the final model. These results are shown in Table 3 and Figure 2.

**Figure 1.**
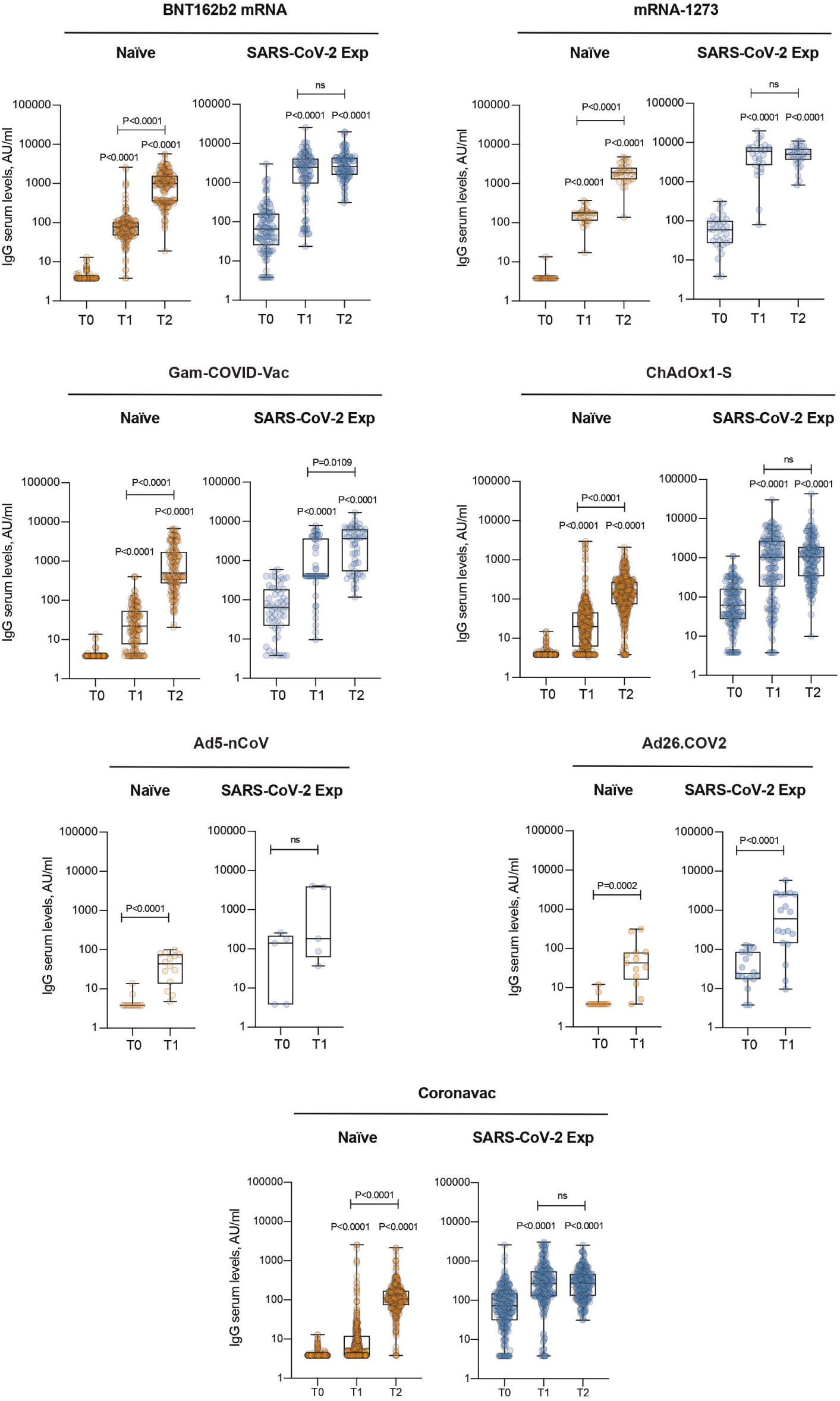
IgG antibody levels by vaccine. IgG antibody response was measured in serum of naïve and SARS-CoV-2 previously exposed (SARS-CoV-2 Exp) subjects at different time points (T0, T1 and T2) and vaccinated with different vaccine types. Samples ≥ 15 AU/mL were considered positive. Log scale on y axis. The box plots show the interquartile range, the horizontal lines show the median values, and the whiskers indicate the minimum-to-maximum range. Each dot corresponds to an individual subject. P-values were determined using 2-tailed Kruskal-Wallis test with Dunn’s multiple comparisons test. P values refer to baseline (T0) when there are no connecting lines.

**Table 2.** Antibody differences between vaccine types

|  | SARS CoV-2 Naïve |  |  |  |  |  |  | p-value |
| --- | --- | --- | --- | --- | --- | --- | --- | --- |
| Vaccine Median (IQR) | BNT162b2 mRNA (n=289) | mRNA-1273 (n=65) | Gam-COVID-Vac(n=213) | Coronavac ((n=582) | ChAdOx1-S (n=666) | Ad5-nCoV Cansino (n=19) | Ad26.COV2(n=31) |  |
| Basal | 3.8(0) | 3.8 (26.6) | 3.8 (0) | 3.8(0.9) | 3.8 (0) | 3.8 (0) | 3.8 (13.5) | p<0.005 |
| After First Dose | 78.1 (55.0) | 175.5 (76.7) | 22.1(44.5) | 5.59 (8.0) | 19.9 (39.6) | 43.45 (55.9) | 42.6 (57.5) | p<0.001 |
| After Second Dose | 998 (1241.0) | 1875 (1190.0) | 501 (1453.7) | 109(97.6) | 140.5 (199.5) | NA | NA | p<0.001 |
|  | SARS CoV-2 Positive |  |  |  |  |  |  |  |
| Basal | 66.9(130.8) | 59.3 (67.9) | 59.8 (157.3) | 72.4 (119.0) | 61.9 (134.8) | 141.0 (175.2) | 24 (66.7) | p<0.001 |
| After First dose | 2500.0(3080.0) | 5920.0 (4705.0) | 400.0 (3252.0) | 264.5 (422.7) | 1025.0 (2489.0) | 182.0 (3734.6) | 606.5 (23.69.0) | p<0.001 |
| After Second Dose | 2630.0 (2672.5) | 4950.0 (3060) | 3620 (5356) | 279(337) | 1020(1538) | NA | NA | p<0.001 |

\*Kruskal-Wallis test was performed for comparison, p<0.05 was considered statistically significant

**Table 3:**
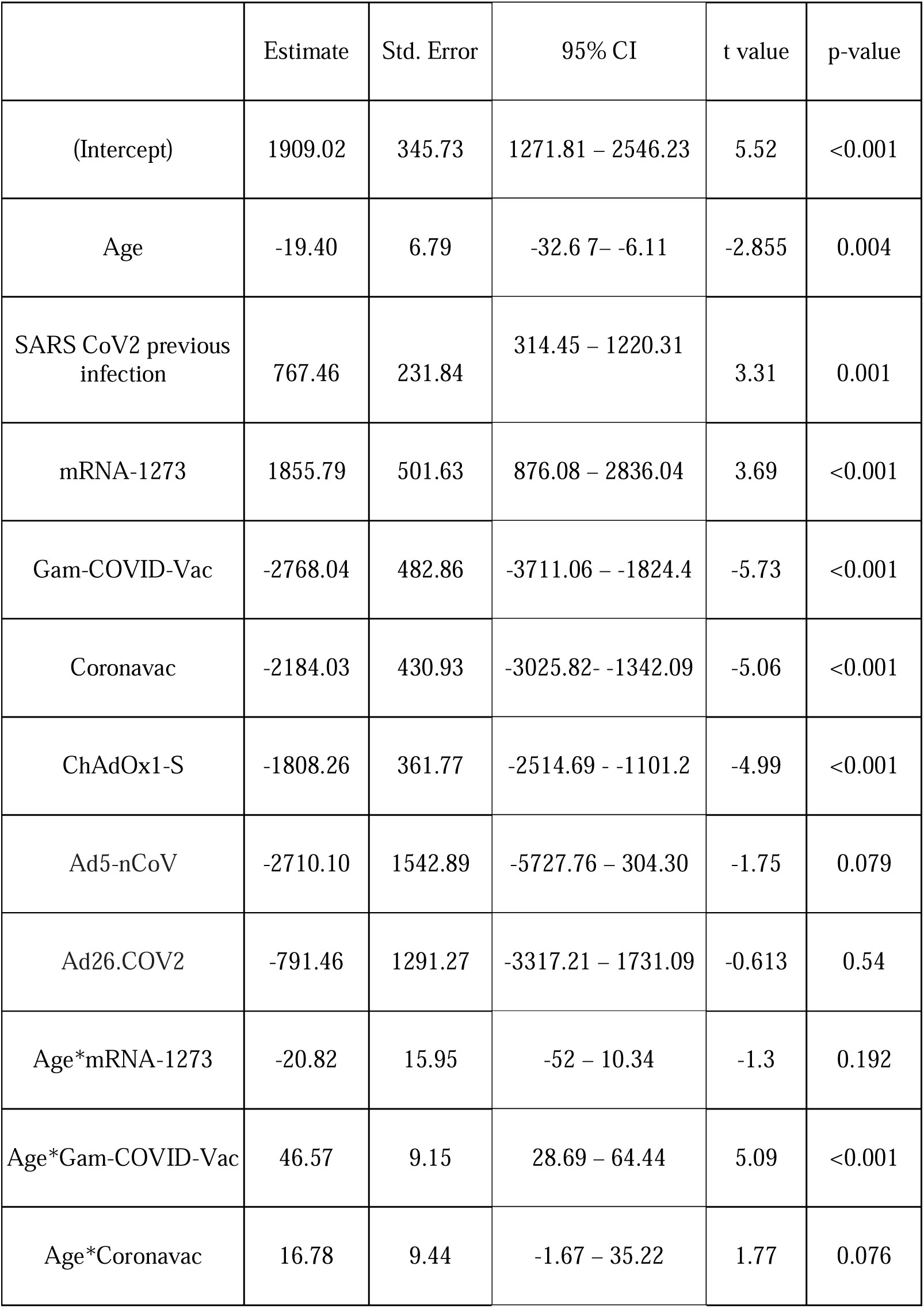
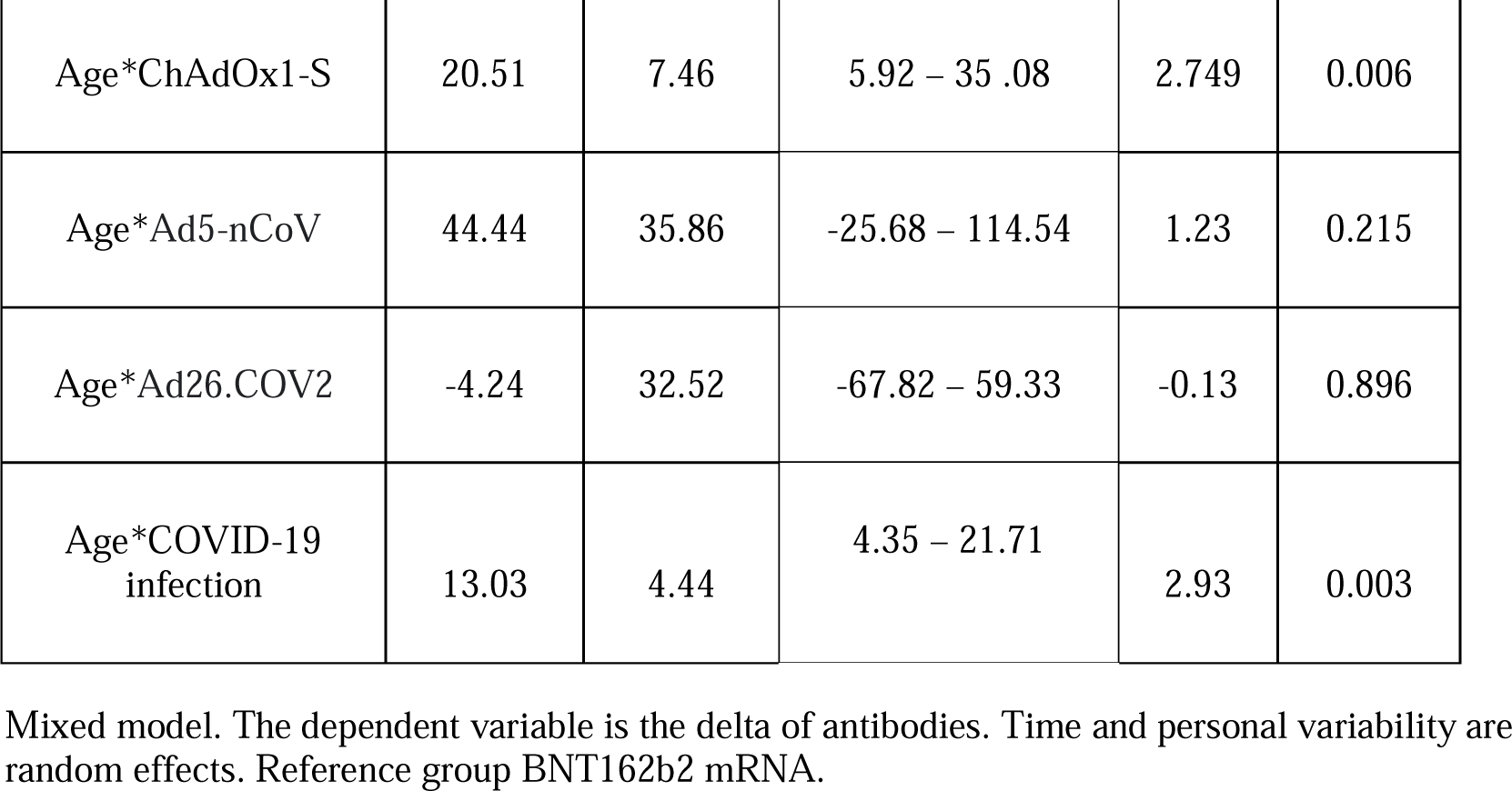
Mixed model of antibody changes

**Figure 2.**
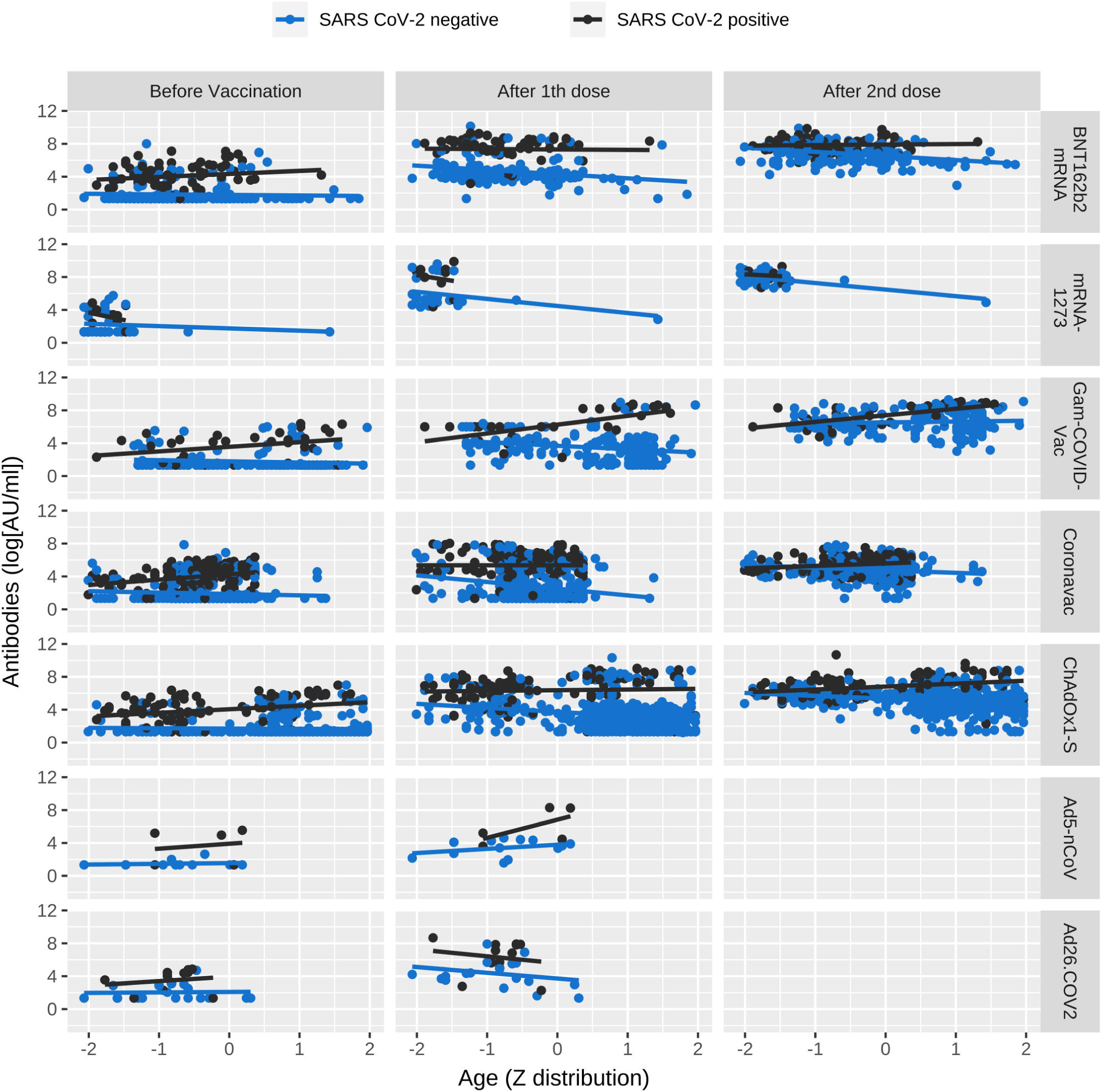
Antibody titers classified by age type of vaccine and Time Figure shows change in antibody concentration through age, divided SARS-CoV-2 history, and vaccine type. Antibodies are expressed in logarithm and age has a Z distribution.

### Adverse Events Following Immunization (AEFI)

At least one AEFI was observed after the first dose in 71% of respondents receiving BNT162b2 mRNA, in 93% with mRNA-1273, 38% with Gam-COVID-Vac, 42% with Coronavac, 51% with ChAdOx1-S, 74% with Ad5-nCoV, and 81% with Ad26.COV2. After the second dose, 65%, 88%, 23%, 33%, and 23% of the respondents experienced at least one AEFI after receiving BNT162b2 mRNA, mRNA-1273, Gam-COVID-Vac, Coronavac, and ChAdOx1-S, respectively. For each vaccine, the majority of the adverse events occurred during the first 24 hrs after injection, either after the first or the second dose. Patients receiving BNT162b2 mRNA, mRNA-1273, Gam-COVID-Vac, Coronavac, ChAdOx1-S, Ad5-nCoV, and Ad26.COV2 subjectively qualified the AEFI after the first dose as “very mild” or “mild” in 85%, 80%, 95%, 84%, 67%, 93%, and 57% of cases, respectively. The AEFI after the second dose were qualified as “very mild” or “mild” by 82%, 49%, 98%, 89%, and 76% of patients receiving BNT162b2 mRNA, mRNA-1273, Gam-COVID-Vac, Coronavac, and ChAdOx1-S respectively. Finally, 49% of patients receiving mRNA-1273 qualified for their adverse events after the second dose as “moderate”. Figure 3 shows the percentages of AEs per vaccine. AEs were evaluated by using PGLM. The model for naïve SARS-CoV-2 subjects showed a negative association of the count of adverse events with age (B=-0.0327, p<0.001) and sex (B=-0.8145, p<0.001). A negative effect with respect to BNT162b mRNA was observed for Gam-COVID-Vac (B=-1.8582, p<0.001), Coronavac (B=-0.5736, p=0.047) and ChAdOx1-S (B=-0.7871, p=0.006). Finally, a positive interaction was found between the delta of antibodies due to vaccination and Gam-COVID-Vac (B=0.5604, p=0.002).

**Figure 3.**
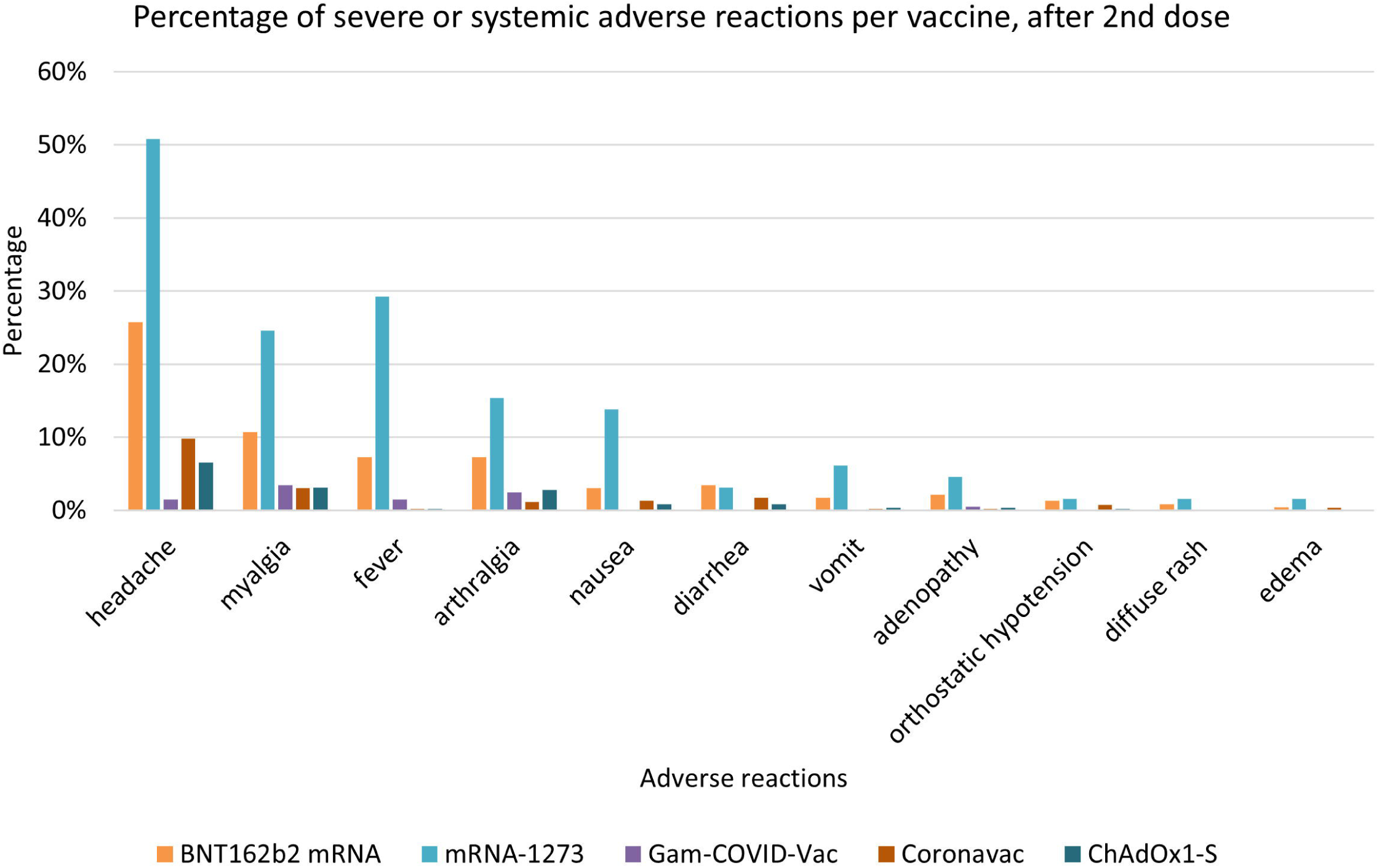
Percentage of systemic or severe adverse events after second dose, stratified by vaccine Graph shows the percentage of systemic or severe adverse events grouped by frequency of appearance

The model for subjects previously exposed to SARS-CoV-2 showed a negative association of the count of adverse events with age (B=-0.0190, p=0.013) and sex (B=-1.0042, p<0.001). The delta of the antibodies showed a positive association with the count of adverse events (B=0.1018, p<0.001). A positive effect with respect to BNT162b mRNA was observed for Gam-COVID-Vac (B=1.2737, p=0.038), whereas a negative effect was observed for Coronavac (B=-1.1304, p<0.001). Negative interactions were also found between the delta of antibodies due to vaccination and the vaccines Gam-COVID-Vac (B=-8.5758, p<0.001) and ChAdOx1-S (B=-1.2386, p=0.013). See Tables 4 and 5 and Figure 4 for details.

**Table 4:** Number of systemic or severe adverse events following vaccination in SARS-CoV-2 Naïve subjects.

| | $\beta$ | $e$ | 95% CI | p-value |
| --- | --- | --- | --- | --- |
| Intercept | 0.73 | 2.08 | 0.95-4.57 | 0.066 |
| Age | -0.03 | 0.96 | 0.95-0.97 | <0.001 |
| Sex (M) | -0.81 | 0.44 | 0.33-0.58 | <0.001 |
| Body Mass Index | 0.015 | 1.01 | 0.99-1.03 | 0.175 |
| Delta IgG | -0.21 | 0.80 | 0.60-1.06 | 0.128 |
| mRNA-1273 | 0.46 | 1.59 | 0.71-3.54 | 0.255 |
| Gam-COVID-Vac | -1.85 | 0.15 | 0.06-0.35 | <0.001 |
| Coronavac | -0.57 | 0.56 | 0.31-0.99 | 0.047 |
| ChAdOx1-S | -0.78 | 0.45 | 0.25-0.79 | 0.006 |
| Delta IgG*mRNA-1273 | 0.13 | 1.14 | 0.78-1.68 | 0.479 |
| Delta IgG*Gam-COVID-Vac | 0.56 | 1.75 | 1.22-2.49 | 0.002 |
| Delta IgG*Coronavac | -1.63 | 0.19 | 0.01-2.39 | 0.201 |
| Delta IgG*ChAdOx1-S | 0.97 | 2.64 | 0.93-7.44 | 0.066 |

Poisson Generalized Linear Model regression. The reference group is BNT162b2 mRNA

**Table 5.**
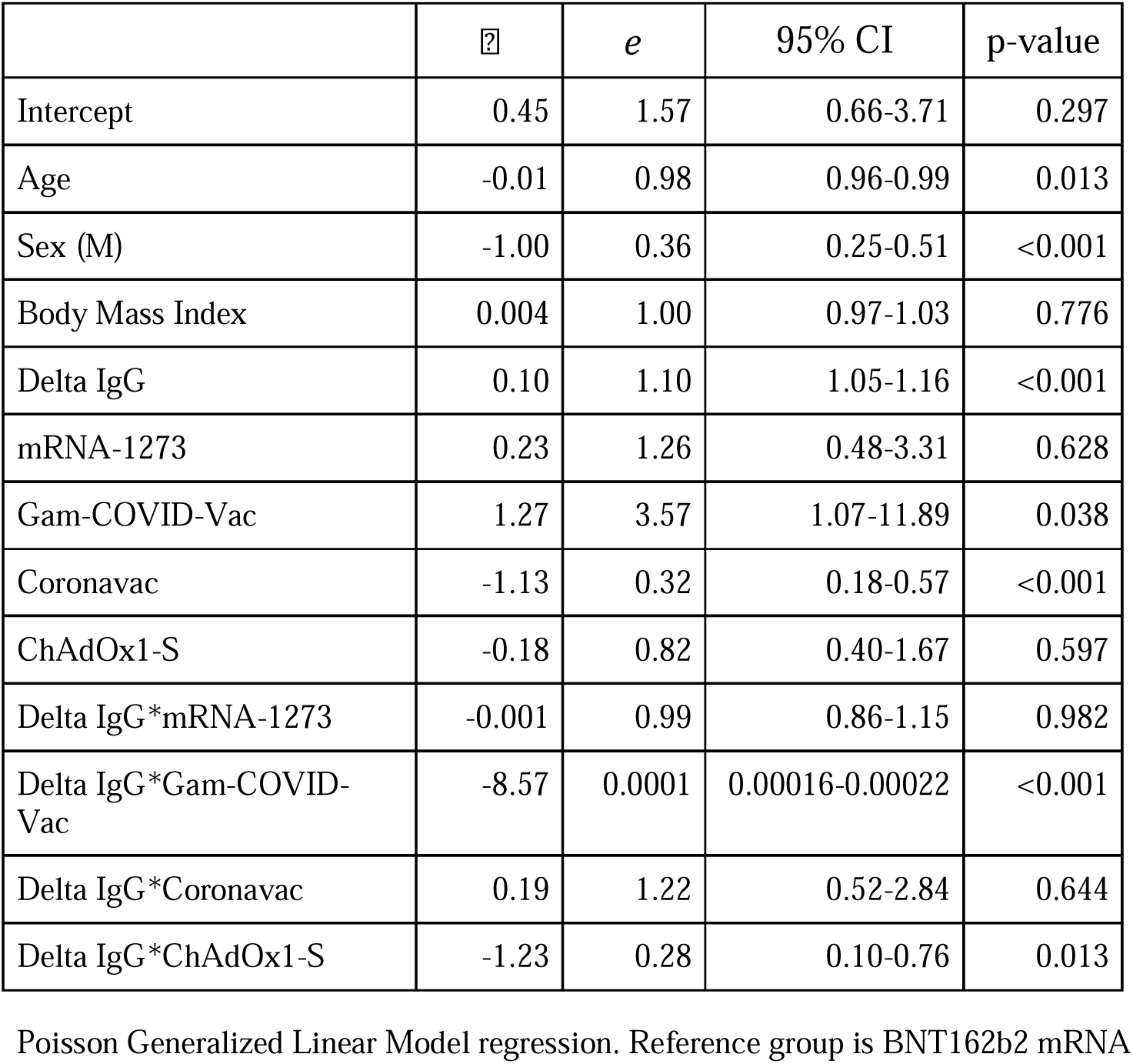
Number of systemic or severe adverse events following vaccination in SARS-CoV-2 previously exposed subjects.

**Figure 4.**
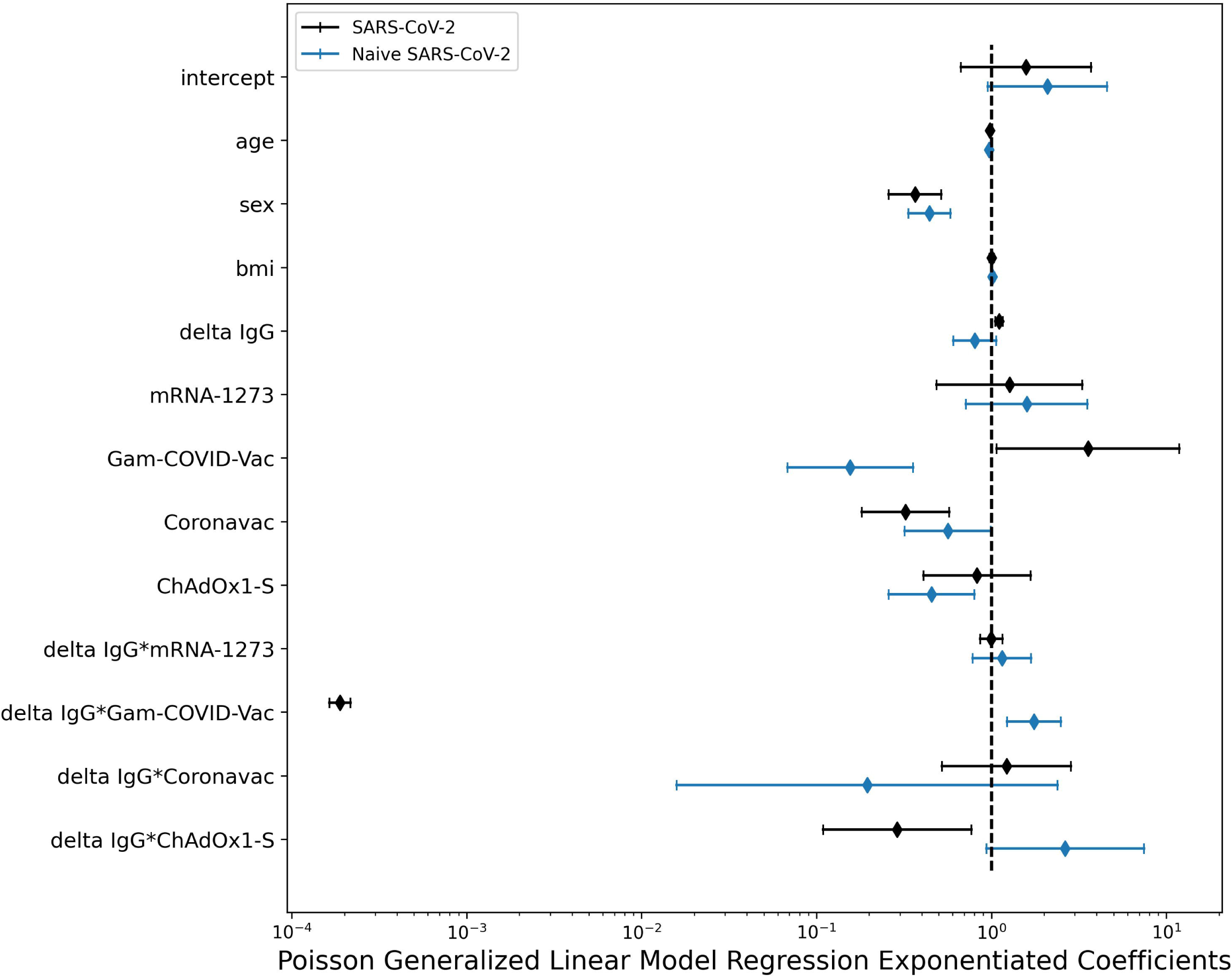
Forest plot for the GLM Poisson model coefficients stratified by SARS-CoV-2 history. Women and young individuals developed more AEs. There is no effect on Body Mass Index. In the naïve SARS-CoV-2 cohort, the vaccines Gam-COVID-Vac, Coronavac and ChAdOx1-S are related to fewer AEFIs after the second dose than BNT162b mRNA. Also, for naïve SARS-CoV-2 patients who received Gam-COVID-Vac, higher antibody levels after the second dose were related to a greater number of AEs. In people previously exposed to SARS-CoV-2, those receiving Gam-COVID-Vac showed a greater number of AEs compared to BNT162b mRNA, while subjects receiving Coronavac showed significantly fewer events when compared to BNT162b mRNA.

### SARS-CoV-2 infection

Subjects were followed up to 28 days after the second dose administration. Between the first and second dose, one (0.34%) patient with BNT162b2 mRNA, 0 with mRNA-1273, 1 (0.46%) with Gam-COVID-Vac, 3 (0.51%) with Coronavac, 4 (0.6%) with ChAdOx1-S, 2 (10%) with Ad5-nCoV and 1 (3%) with Ad26.COV2 got the SARS-CoV-2 infection. After the second dose, 4 (1.3%) with BNT162b2 mRNA, 1 (1.5%) with mRNA-1273, 0 with Gam-COVID-Vac, 18 (3%) with Coronavac, and 11 (1.6%) with ChAdOx1-S became infected with SARS-CoV-2 (p<0.01).

## Discussion

This multicentric study compared the 21-28 day seroconversion, the AEFI after first and second doses, and the associated predictors of AEs of seven of the most common vaccines used worldwide: BNT162b2 mRNA, mRNA-1273, Gam-COVID-Vac, Coronavac, ChAdOx1-S, Ad5-nCoV, and Ad26.COV2(4).

All vaccines showed immunogenicity and seroconversion after the complete vaccination scheme. The greatest change in antibody levels occurred with mRNA-1273, followed by BNT162b2 mRNA and Gam-COVID-Vac. Previous studies have shown a greater change in antibody titers for mRNA vector vaccines(12–14).

Our study found that the previous history of SARS-CoV-2 was related to an increase in change in antibody levels, as confirmed elsewhere (12). Also, we found higher levels in antibodies in older patients when they had previous SARS-CoV-2 infection and received Gam-COVID Vac or ChAdOx1-S. The previous study from Brazil did not find any association of antibody levels with age. We also found that except for the Gam-COVID Vac, in the other vaccines requiring two doses, the first dose was sufficient to induce maximal antibody levels in subjects with a history of SARS-CoV-2, as previously shown for the BNT162b2 mRNA vaccine(9,15–19)

The study of Logunov et al. that tested the efficacy of Gam-COVID Vac reported a higher increase of anti-SARS-CoV-2 IgG levels in the group of 18-30 years and no particular change in other age groups(20); however they excluded patients with previous SARS-CoV-2 infection, so these results are not comparable.

In the study conducted by Ramasamy et al., similar antibody titers were seen 28 days after the boost vaccination of ChAdOx1 across all groups, regardless of age or vaccine dose 21. It is possible that older patients in our group of the study had a more severe disease during SARS-CoV-2 infection and that they developed higher antibody titers, as it has been reported that patients aged higher than 60 can develop this condition (21).

For most patients, at least one AEFI occurred with all types of vaccines during the first 24 hrs. after injection, and most of them qualified the symptoms as “very mild” or “mild” both after the first and second dose, except for mRNA-1273 for which nearly half of the volunteers reported moderate symptoms after the second dose as reported previously(18,20,22–27).

Our models showed a higher risk for women and young people to experience AEs. A real-world study that reported AEFI of patients receiving BNT162b2 mRNA, mRNA-1273, or Ad26COV2 in one or two doses (28), an independent studies conducted with Gam-COVID also confirmed this results (26,29). The study of Ramasamy et al. on ChAdOx1 showed more adverse events in the group of 18-55 years old(22). The study conducted by Scott A et al. showed that severe adverse events (grade 5) were more frequent in women with Ad5-nCoV (25).

On one hand, for the Naïve SARS-CoV-2 cohort, the vaccines Gam-COVID-Vac, Coronavac, and ChAdOx1-S were related to fewer AEs after the second dose than BNT162b mRNA, and to the best of our knowledge, there are no studies that compare AEs across these types of vaccines. Also, the delta of antibodies was related to AEs after the second dose in Gam-COVID-Vac. As previously published, mRNA-1273 had higher AEFIs when compared to BNT162b2 mRN(30).

On the other hand, for subjects previously exposed to SARS-CoV-2, those receiving Gam-COVID-Vac showed a greater number of AEs than BNT162b mRNA, while subjects receiving Coronavac showed significantly fewer events when compared to BNT162b mRNA. In general, an increase in the antibody levels was related to an increase in the number of AEFIs, but for people receiving Gam-COVID-Vac or ChAdOx1-S, greater changes in antibody levels corresponded to fewer AEs. This last result should be taken with caution due to the limited number of patients falling in the category. To the best of our knowledge, this has not been previously shown. The study conducted by Sung Hee-Lim et al. that compared the antibody response with adverse events in patients vaccinated with BNT162b mRNA or ChAdOx1-S reported no association with antibody change and adverse events; however, patients previously infected with SARS-CoV-2 were excluded (31).

This study shows a low rate of infection in all vaccine groups. However, the highest rate of SARS-CoV-2 infection in this follow-up period was for the one-dose vaccines Ad5-nCoV and Ad26.COV2. Also, Coronavac had a high rate of infection. It is important to consider that the patients were followed for a short period.

As a limitation of our study, we recognize that we had a small sample size in some vaccine groups such as Ad5-nCoV; however, to the best of our knowledge, there is no other study where this type of vaccine has been compared to other types of vaccines in a real-world setting. It will be of interest to follow the IgG titers for a longer period and to evaluate the effect of heterologous combinations of these vaccines.

This comparative study of vaccine types shows positive immunogenicity and seroconversion of BNT162b2 mRNA, mRNA-1273, Gam-COVID-Vac, Coronavac, ChAdOx1-S, Ad5-nCoV, and Ad26.COV2. The highest IgG response was for the mRNA vector vaccines and the lower for the inactivated vaccine. Women and young individuals developed more AEs. In the naïve SARS-CoV-2 cohort, the vaccines Gam-COVID-Vac, Coronavac, and ChAdOx1-S were related to fewer AEs after the second dose of BNT162b mRNA. For patients who received Gam-COVID-Vac, higher antibody levels after the second dose were related to a greater number of AEs. In people previously exposed to SARS-CoV-2, those receiving Gam-COVID-Vac showed a greater number of AEs than BNT162b mRNA, while subjects receiving Coronavac showed significantly fewer events when compared to BNT162b mRNA. In general, an increase in antibody levels was related to AEs.

## Data Availability

All data produced in the present study are available upon reasonable request to the authors

## Abbreviations

(AEFI): Adverse events following immunization
(AEs): Severe adverse events

## Data Availability Statement

Data are available upon reasonable request to the authors.

## Ethics statement

Ethics committee/local Institutional Review Board from Hospital Clinica Nova, Humanitas Clinical and Research Center, Fundacion San Francisco Xavier, Ternium Health Center in Rio, Hospital Municipal San Jose, Hospital Interzonal de Agudos San Felipe gave ethical approval.

## Author Approval

all authors all authors read an approved the final version of the manuscript.

## Author Contributions

Conceptualization: MERI, MR, MT, MASS, and EA. Formal analysis: MERI, MR, MT, MASS, AGC, CP, RL, S, MM, and RB. Investigation: MERI, AGC, MR, MT, MASS, CP, RL, RS, RB, MM, DRS, YGHR, AGAV, GFDRP, IABF, RGF, EZ, ARAD, MHG, ABB, CA, GPB, MT, and MR. Resources: MR, MT, and MASS Writing – original draft: MERI, AGC, CP, MM, RS, RL, RB, MASS, EA, DRS, YGHR, AGAV, GFDRP, IABF, RGF, EZ, ARAD, MHG, ABB, CA, GPB, MT, MR. Writing – review and editing: MERI, AGC, CP, MM, RS, RL, RB, MASS, EA, DRS, YGHR, AGAV, GFDRP, IABF, RGF, EZ, ARAD, MHG, ABB, CA, GPB, MT, MR. Project administration: MR, MT, MASS. Supervision: MERI, MT, MR, and MASS. Funding acquisition: MT, MR, and MASS. All authors contributed to the article and approved the submitted version.

## Acknowledgments

This project has been supported by the Techint Group of Companies.

We thank Erika Bienek (Techint Group Community Relations Director), Walter Bruno (Humanitas Communications Director), Cesar Bueno (Fundación São Francisco Xavier President), Mario Galli (Techint Group Communications Director), Pablo Sturiale (Apsot Director) for their support of the work and discussions.

## References

1. WHO Director-General’s opening remarks at the media briefing on COVID-19 - 11 March 2020. https://www.who.int/dg/speeches/detail/who-director-general-s-opening-remarks-at-the-media-briefing-on-covid-19---11-march-2020 [Accessed April 19, 2020]

2. Kannan S, Shaik Syed Ali P, Sheeza A, Hemalatha K. COVID-19 (Novel Coronavirus 2019) – recent trends. European Review for Medical and Pharmacological Sciences (2020) 24:2006– 2011. doi: 10.26355/eurrev_202002_20378

3. Kumar S, Nyodu R, Maurya VK, Saxena SK. “Morphology, Genome Organization, Replication, and Pathogenesis of Severe Acute Respiratory Syndrome Coronavirus 2 (SARS-CoV-2).,” In: Saxena SK, editor. Coronavirus Disease 2019 (COVID-19). Medical Virology: From Pathogenesis to Disease Control. Singapore: Springer Singapore (2020). p. 23–31 doi: 10.1007/978-981-15-4814-7_3

4. Covid-19 Vaccine Tracker: Latest Updates - The New York Times. https://www.nytimes.com/interactive/2020/science/coronavirus-vaccine-tracker.html [Accessed January 11, 2022]

5. Mascellino MT, Di Timoteo F, De Angelis M, Oliva A. Overview of the Main Anti-SARS-CoV-2 Vaccines: Mechanism of Action, Efficacy and Safety. IDR (2021) Volume 14:3459–3476. doi: 10.2147/IDR.S315727

6. von Elm E, Altman DG, Egger M, Pocock SJ, Gøtzsche PC, Vandenbroucke JP, STROBE Initiative. The Strengthening the Reporting of Observational Studies in Epidemiology (STROBE) statement: guidelines for reporting observational studies. Ann Intern Med (2007) 147:573–577. doi: 10.7326/0003-4819-147-8-200710160-00010

7. Chen M, Yuan Y, Zhou Y, Deng Z, Zhao J, Feng F, Zou H, Sun C. Safety of SARS-CoV-2 vaccines: a systematic review and meta-analysis of randomized controlled trials. Infect Dis Poverty (2021) 10:94. doi: 10.1186/s40249-021-00878-5

8. Bonelli F, Sarasini A, Zierold C, Calleri M, Bonetti A, Vismara C, Blocki FA, Pallavicini L, Chinali A, Campisi D, et al. Clinical and Analytical Performance of an Automated Serological Test That Identifies S1/S2-Neutralizing IgG in COVID-19 Patients Semiquantitatively. J Clin Microbiol (2020) 58: doi: 10.1128/JCM.01224-20

9. Levi R, Azzolini E, Pozzi C, Ubaldi L, Lagioia M, Mantovani A, Rescigno M. One dose of SARS-CoV-2 vaccine exponentially increases antibodies in individuals who have recovered from symptomatic COVID-19. Journal of Clinical Investigation (2021) 131:e149154. doi: 10.1172/JCI149154

10. Levi R, Ubaldi L, Pozzi C, Angelotti G, Sandri MT, Azzolini E, Salvatici M, Savevski V, Mantovani A, Rescigno M. The antibody response to SARS-CoV-2 infection persists over at least 8 months in symptomatic patients. Commun Med (2021) 1:32. doi: 10.1038/s43856-021-00032-0

11. Sandri MT, Azzolini E, Torri V, Carloni S, Pozzi C, Salvatici M, Tedeschi M, Castoldi M, Mantovani A, Rescigno M. SARS-CoV-2 serology in 4000 health care and administrative staff across seven sites in Lombardy, Italy. Sci Rep (2021) 11:12312. doi: 10.1038/s41598-021-91773-4

12. Szczepanek J, Skorupa M, Goroncy A, Jarkiewicz-Tretyn J, Wypych A, Sandomierz D, Jarkiewicz-Tretyn A, Dejewska J, Ciechanowska K, Palgan K, et al. Anti-SARS-CoV-2 IgG against the S Protein: A Comparison of BNT162b2, mRNA-1273, ChAdOx1 nCoV-2019 and Ad26.COV2.S Vaccines. Vaccines (2022) 10:99. doi: 10.3390/vaccines10010099

13. Lau O, Vadlamudi NK. Immunogenicity and Safety of the COVID-19 Vaccines Compared With Control in Healthy Adults: A Qualitative and Systematic Review. Value in Health (2021)S1098301521017344. doi: 10.1016/j.jval.2021.09.003

14. Rotshild V, Hirsh-Raccah B, Miskin I, Muszkat M, Matok I. Comparing the clinical efficacy of COVID-19 vaccines: a systematic review and network meta-analysis. Sci Rep (2021) 11:22777. doi: 10.1038/s41598-021-02321-z

15. Krammer F, Srivastava K, the PARIS team, Simon V. Robust spike antibody responses and increased reactogenicity in seropositive individuals after a single dose of SARS-CoV-2 mRNA vaccine. [preprint]. Allergy and Immunology (2021). doi: 10.1101/2021.01.29.21250653

16. Krammer F, Srivastava K, Alshammary H, Amoako AA, Awawda MH, Beach KF, Bermúdez-González MC, Bielak DA, Carreño JM, Chernet RL, et al. Antibody Responses in Seropositive Persons after a Single Dose of SARS-CoV-2 mRNA Vaccine. N Engl J Med (2021) 384:1372– 1374. doi: 10.1056/NEJMc2101667

17. Saadat S, Rikhtegaran Tehrani Z, Logue J, Newman M, Frieman MB, Harris AD, Sajadi MM. Binding and Neutralization Antibody Titers After a Single Vaccine Dose in Health Care Workers Previously Infected With SARS-CoV-2. JAMA (2021) 325:1467–1469. doi: 10.1001/jama.2021.3341

18. Sadoff J, Gray G, Vandebosch A, Cárdenas V, Shukarev G, Grinsztejn B, Goepfert PA, Truyers C, Fennema H, Spiessens B, et al. Safety and Efficacy of Single-Dose Ad26.COV2.S Vaccine against Covid-19. N Engl J Med (2021) 384:2187–2201. doi: 10.1056/NEJMoa2101544

19. Samanovic MI, Cornelius AR, Gray-Gaillard SL, Allen JR, Karmacharya T, Wilson JP, Hyman SW, Tuen M, Koralov SB, Mulligan MJ, et al. Robust immune responses after one dose of BNT162b2 mRNA vaccine dose in SARS-CoV-2 experienced individuals. [preprint]. Infectious Diseases (except HIV/AIDS) (2021). doi: 10.1101/2021.02.07.21251311

20. Logunov DY, Dolzhikova IV, Shcheblyakov DV, Tukhvatulin AI, Zubkova OV, Dzharullaeva AS, Kovyrshina AV, Lubenets NL, Grousova DM, Erokhova AS, et al. Safety and efficacy of an rAd26 and rAd5 vector-based heterologous prime-boost COVID-19 vaccine: an interim analysis of a randomised controlled phase 3 trial in Russia. The Lancet (2021) 397:671–681. doi: 10.1016/S0140-6736(21)00234-8

21. Cannistraci CV, Valsecchi MG, Capua I. Age-sex population adjusted analysis of disease severity in epidemics as a tool to devise public health policies for COVID-19. Sci Rep (2021) 11:11787. doi: 10.1038/s41598-021-89615-4

22. Ramasamy MN, Minassian AM, Ewer KJ, Flaxman AL, Folegatti PM, Owens DR, Voysey M, Aley PK, Angus B, Babbage G, et al. Safety and immunogenicity of ChAdOx1 nCoV-19 vaccine administered in a prime-boost regimen in young and old adults (COV002): a single-blind, randomised, controlled, phase 2/3 trial. The Lancet (2020) 396:1979–1993. doi: 10.1016/S0140-6736(20)32466-1

23. Polack FP, Thomas SJ, Kitchin N, Absalon J, Gurtman A, Lockhart S, Perez JL, Pérez Marc G, Moreira ED, Zerbini C, et al. Safety and Efficacy of the BNT162b2 mRNA Covid-19 Vaccine. N Engl J Med (2020) 383:2603–2615. doi: 10.1056/NEJMoa2034577

24. Baden LR, El Sahly HM, Essink B, Kotloff K, Frey S, Novak R, Diemert D, Spector SA, Rouphael N, Creech CB, et al. Efficacy and Safety of the mRNA-1273 SARS-CoV-2 Vaccine. N Engl J Med (2021) 384:403–416. doi: 10.1056/NEJMoa2035389

25. Halperin SA, Ye L, MacKinnon-Cameron D, Smith B, Cahn PE, Ruiz-Palacios GM, Ikram A, Lanas F, Lourdes Guerrero M, Muñoz Navarro SR, et al. Final efficacy analysis, interim safety analysis, and immunogenicity of a single dose of recombinant novel coronavirus vaccine (adenovirus type 5 vector) in adults 18 years and older: an international, multicentre, randomised, double-blinded, placebo-controlled phase 3 trial. The Lancet (2022) 399:237–248. doi: 10.1016/S0140-6736(21)02753-7

26. Jarynowski A, Semenov A, Kaminski M, Belik V. Mild Adverse Events of Sputnik V Vaccine in Russia: Social Media Content Analysis of Telegram via Deep Learning. J Med Internet Res (2021) 23:e30529. doi: 10.2196/30529

27. Halder A, Imamura H, Condon S, Boroughs K, Nilsson SC, Anderson T, Caterson ID. Pfizer/BioNtech BNT162b2: adverse events and insights from an Australian mass vaccination clinic for COVID-19. Intern Med J (2022) 52:121–124. doi: 10.1111/imj.15623

28. Beatty AL, Peyser ND, Butcher XE, Cocohoba JM, Lin F, Olgin JE, Pletcher MJ, Marcus GM. Analysis of COVID-19 Vaccine Type and Adverse Effects Following Vaccination. JAMA Netw Open (2021) 4:e2140364. doi: 10.1001/jamanetworkopen.2021.40364

29. Babamahmoodi F, Saeedi M, Alizadeh-Navaei R, Hedayatizadeh-Omran A, Mousavi SA, Ovaise G, Kordi S, Akbari Z, Azordeh M, Ahangarkani F, et al. Side effects and Immunogenicity following administration of the Sputnik V COVID-19 vaccine in health care workers in Iran. Sci Rep (2021) 11:21464. doi: 10.1038/s41598-021-00963-7

30. Meo SA, Bukhari IA, Akram J, Meo AS, Klonoff DC. COVID-19 vaccines: comparison of biological, pharmacological characteristics and adverse effects of Pfizer/BioNTech and Moderna Vaccines. Eur Rev Med Pharmacol Sci (2021) 25:1663–1669. doi: 10.26355/eurrev_202102_24877

31. Lim S-H, Choi S-H, Kim B, Kim J-Y, Ji Y-S, Kim S-H, Kim C-K, Kim T, Choo E-J, Moon J-E, et al. Serum Antibody Response Comparison and Adverse Reaction Analysis in Healthcare Workers Vaccinated with the BNT162b2 or ChAdOx1 COVID-19 Vaccine. Vaccines (2021) 9:1379. doi: 10.3390/vaccines9121379

