## Supplemental information for "Analysis of immunization time, amplitude, and adverse events of seven different vaccines against SARS-CoV-2 across four different countries"

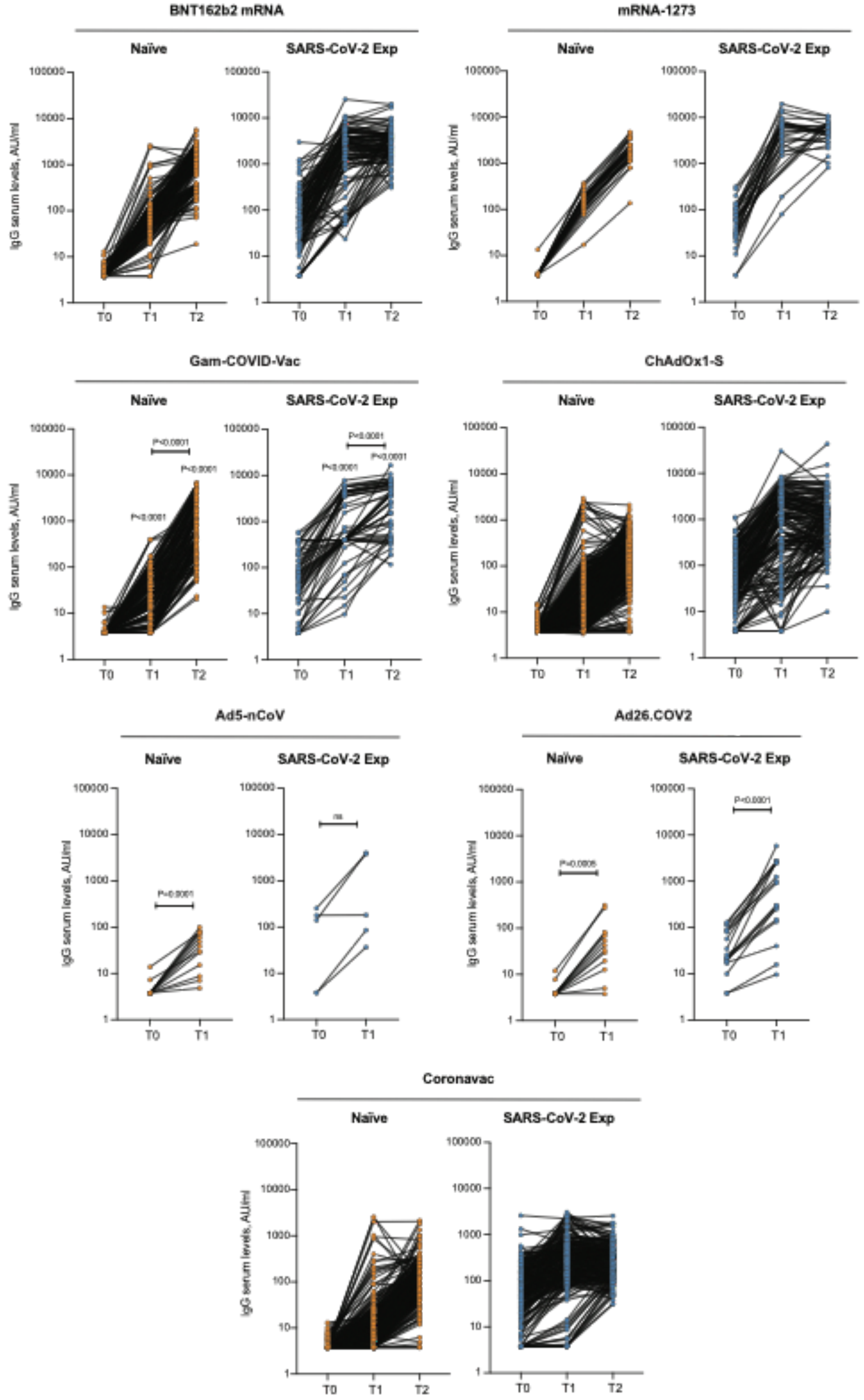


**Figure S1. Kinetics of antibody response.**

IgG antibody response was measured in serum of naïve and SARS-CoV-2 previously exposed (SARS-CoV-2 Exp) subjects at different time points (T0, T1 and T2) and vaccinated with different vaccine types. Samples ≥ 15 AU/mL were considered positive. Log scale on y axis. Spaghetti plots showing the trends for each individual subject by linked dots. P values were determined using Friedman test with Dunn's multiple comparisons test. P values refer to baseline (T0) when there are no connecting lines.
